# Variant-Specific Landscape of Mutual Exclusivity Among BRAF, EGFR, and KRAS Oncogenes in Human Cancer

**DOI:** 10.1101/2023.10.21.23297089

**Authors:** Freya Vaeyens, Jan-Patrick Hetzel, Marco Mernberger, Carolien Eggermont, Catharina Olsen, Ken Maes, Jelle Vlaeminck, Frederik Hes, Martin Pichler, Philippe Giron, Oleg Timofeev, Maxim Noeparast

## Abstract

In this cross-sectional study, we report the findings of our investigation into the mutual exclusivity (ME) and co-occurrence (CO) patterns of BRAF, KRAS, and EGFR mutations in human cancer. Our analysis acknowledges previously overlooked mutational subtypes with distinct clinical implications. Creating an automated R framework, we analyzed mutation data from 64807 unique cBioPortal samples, 1570 cell lines, and 2714 unique Belgian cancer samples. Consistently, across all three datasets, we observe that co-occurrence is less likely among class I BRAF, Hydrolysis KRAS, and Classical-like EGFR mutations. Bilateral variant-assigned CO matrices uncover novel inter-class and inter-type CO and ME scenarios, encompassing conventional and atypical mutations. Besides Class I BRAF, various mutation classes exhibit diverse CO patterns, justifying the need to refine mutational classifications. We provide a variant-specific database for precision oncology showcasing ME among three actionable oncogenes. These findings may guide the discovery of novel synthetically lethal interactions for targeted cancer therapy.

## Introduction

At least 40% of human cancers are associated with aberrant activity of the ERK^♠^ (**E**xtracellular signal-**R**egulated **K**inase) pathway^1,2^. Three frequently mutated effectors of the ERK pathway in cancer, namely BRAF, KRAS, and EGFR, are clinically actionable^3,4^ or being explored^5^. Mutual exclusivity (ME) and co-occurrence (CO) patterns among mutations of these oncogenes have been studied, often without acknowledging the mutational classes concomitantly ^2,6,7^. The consensus is that BRAF∩KRAS and EGFR∩KRAS mutation pairs typically do not co-exist^8–11^. Of note, targeted therapeutics are virtually mutation-specific and exert differential activity towards distinctive variants, and various mutants of the same gene can have distinctive pathway dependencies^3–5,12–18^. We have learned from several years of mutant-BRAF, -KRAS, and -EGFR therapy that mutation-specific targeting is more efficient than merely targeting the pathway dependencies^19^. Moreover, in recent years, mutually exclusive mutational scenarios have provided ground for discovering synthetically lethal targets^20^. As such, spotting the very variants in mutually exclusive or co-occurring scenarios is of high translational significance.

Herein, we conducted a cross-sectional data analysis study by consulting a publicly available cancer database cBioPortal to address the mutual exclusivity or co-occurrence of BRAF, KRAS, and EGFR mutations in pairs while acknowledging these genes’ mutational classes and mutation types. We corroborated our primary findings in a cancer cell line database. Finally, we conducted an analogous analysis on an original and manually curated dataset of Belgian cancer patients to validate the findings from our initial analysis.

## Results

### Discovery of BRAF, KRAS, and EGFR mutation types with Mutually Exclusive or Co-occurring tendencies

In this study, we distinguished three BRAF classes^3^, three KRAS classes^5^, and four EGFR classes^4^ (full description in Table 1). We limited our analyses first to mutations and excluded structural variants and copy number alterations. To gain an updated and comprehensive insight about BRAF, KRAS, and EGFR mutation classes in human cancer, we measured class frequencies for each gene within patients and cell lines datasets (Fig. 1a). Next, we determined mutation counts against their relative rank in patients (Fig. 1b). Interestingly, we found the Sorafenib-senstitive^21^ (anecdotal) BRAF^G469R^, which is predicted to belong to class II, to be among the top ten recurring BRAF mutations. Also, two atypical EGFR variants, including Icotinib^22^- and mAb806-sensitive^23^ EGFR^A289V^ and Dacomitinib-sensitive^24^ (anecdotal) EGFR^G598V^, appeared among the top ten frequent EGFR variants.

**Fig. 1:**
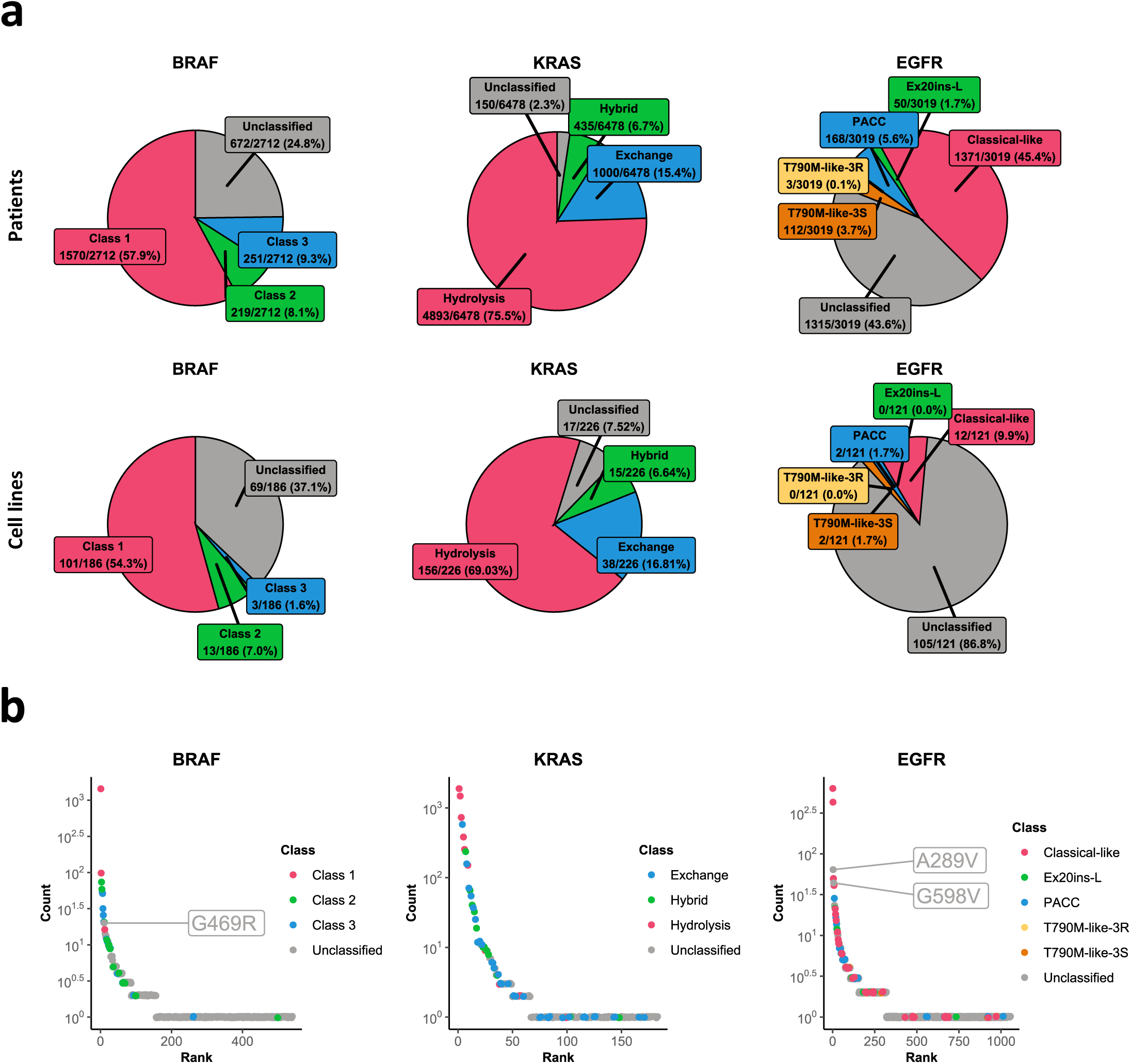
Frequency of Mutational classes and Mutational types in human cancer. (**a**) Absolute and relative frequency of BRAF, KRAS, and EGFR mutational classes in human cancer (as described in Table 1) among patient samples (top) and cell lines (bottom). Different classes of each gene are color-coded and will appear accordingly in the following figures. We queried data of 68479 samples from 64911 cancer patients, encompassing 2710, 6442 and 2999 samples with BRAF, KRAS, and EGFR mutations, respectively. Duplicate patient data were removed as described in the methods section, and concerning multiple samples of the same patient, only the earliest sample was considered. For cell lines, 1570 cell lines in Cancer Cell Line Encyclopedia were queried, and those with BRAF, KRAS, or EGFR mutations were considered for this analysis. (**b**) Scatter plots show the absolute count of each variant against its frequency rank for BRAF (left), KRAS (middle), and EGFR (right) mutations while assigned to their respective class. Lower ranks indicate higher prevalence in human cancer.

**Table. 1:** Classification of BRAF, KRAS, and EGFR mutations acknowledged in this study. According to a widely accepted classification^3^, BRAF mutations are classified into three classes. Class I variants, which include the typical BRAF^V600E^, are known to function as constitutively active monomers and can signal independent of upstream RAS or another RAF isoform CRAF. Atypical BRAF mutations belong to either class II or class III. Class II is suggested to activate the ERK pathway independent of RAS and CRAF but signals as homodimers and activates the ERK pathway at lower levels than class I. BRAF^G469A^ is the well-studied class II BRAF in cancer, which activates ERK slightly less than BRAF^V600E 40^. Class III comprises the kinase-impaired BRAF mutants, which activate the ERK pathway via allosteric transactivation of the CRAF in a RAS-dependent manner^40,41^. For KRAS mutations, we considered the most updated classification described by Johnson et al.^5^. Concerning cancer-related KRAS mutations, three classes are defined according to the impact of mutations on two critical features of the KRAS protein. KRAS can hydrolyze GTP and oscillate back to inactive conformation^5^. Mutations that lead to loss of GTP-hydrolyzing function are classified as class I (Hydrolysis)^5^. Moreover, Guanine Nucleotide Exchange factors (GEFs) and corresponding scaffolding proteins enable KRAS to dislodge GDP, accept GTP, and transit to the active conformation^5^. Mutations that lead to the gain of the KRAS Exchange function are classified as class II (Exchange). Furthermore, the third class comprises the KRAS mutations that affect both the Hydrolysis and Exchange (class III: Hybrid)^5^. Hydrolysis KRAS mutations are the most prevalent KRAS variants in human cancer^5^. With regards to EGFR mutations, we considered a recent classification described by Robichaux et.al^4^ which is based on the structural impact of mutations on EGFR protein and, in particular, EGFR’s drug-binding pocket (DBP) as well as the consequences of mutations on drug response. Accordingly, EGFR mutations are classified into four groups. Classical-likes are those mutations positioned relatively far from DBP and have no or inconsiderable effect on EGFR affinity for the three generations of the Tyrosine Kinase Inhibitors (TKIs)^4^. T790M-like class includes those EGFR variants with at least one mutation in the hydrophobic core of the EGFR ATP binding pocket, at the gatekeeper residue alone or with other mutations^4^. T790M-like mutations increase the affinity of EGFR for ATP and present two sub-groups that we distinguished in our analyses, namely T790M-like-3R and T790M-like-3S; the latter can be sensitive to 3^rd^ generation TKIs. Exon 20 loop insertions (Ex20ins-L) involve the C-terminal loop of the αC-helix and induce a global impact on EGFR DBP by altering both P-loop (inward surface of the ATP binding pocket) and αC-helix conformations. They indirectly affect the affinity for inhibitors. These mutations exhibit differential sensitivity to TKIs^4^. The P-loop αC-helix compressing (PACC) represents mutations close to DBP. Their conformational impact, less than Ex20ins variants, affects either the P-loop or αC-helix alone or concomitantly^4^. They directly or indirectly (like Ex20ins-L) influence the drug binding. These mutations might sensitize to 2^nd^ generation TKIs. Note that our dataset was minimal with Ex20ins and PACC variants. Therefore, we did not distinguish corresponding sub-groups (for detailed and more comprehensive descriptions of EGFR classes, refer to Robichaux et.al^4^).

|  |  | Examples | Basis of classification |
| --- | --- | --- | --- |
| **BRAF class** | I | V600E/K/R (three members only) | (In)dependency on dimerization/RAS+CRAF for signaling |
|  | II | G469A/V, L597V |  |
|  | III | D594A/D/E, G466A |  |
| **KRAS class** | Hydrolysis | G12C/D/V | Loss of GTPase activity, a gain in Guanosin Exchange Factor recruitment, or a combination of both |
|  | Exchange | G13D, V14I, A18T |  |
|  | Hybrid | A59T, G60D, Q61H |  |
| **EGFR class** | Classical-like | Ex19del, L858R, L861Q | Mutation impact on EGFR drug-binding domain 3D structure and drug response |
|  | Ex20ins-L | A767insASV, D770insNPG, H773insNPH |  |
|  | PACC | (Ex19del & L792H), (L858R & C797S), L718V |  |
|  | T790M-like-3R | (Ex19del & T790M & C797S), (L858R & T790M & L718Q), (L718Q & T790M) |  |
|  | T790M-like-3S | T790M, (Ex19del & T790M & L718V), (L858R & T790M) |  |

Next, we conducted two-sided Fisher’s exact tests to assess whether the proportion of each class differs when comparing groups. We identified under- or over-represented classes in the CO groups (Table 2a). To have an overview of single variants, we also broke down our analysis into each variant assigned to its class (Fig. 2).

**Fig. 2:**
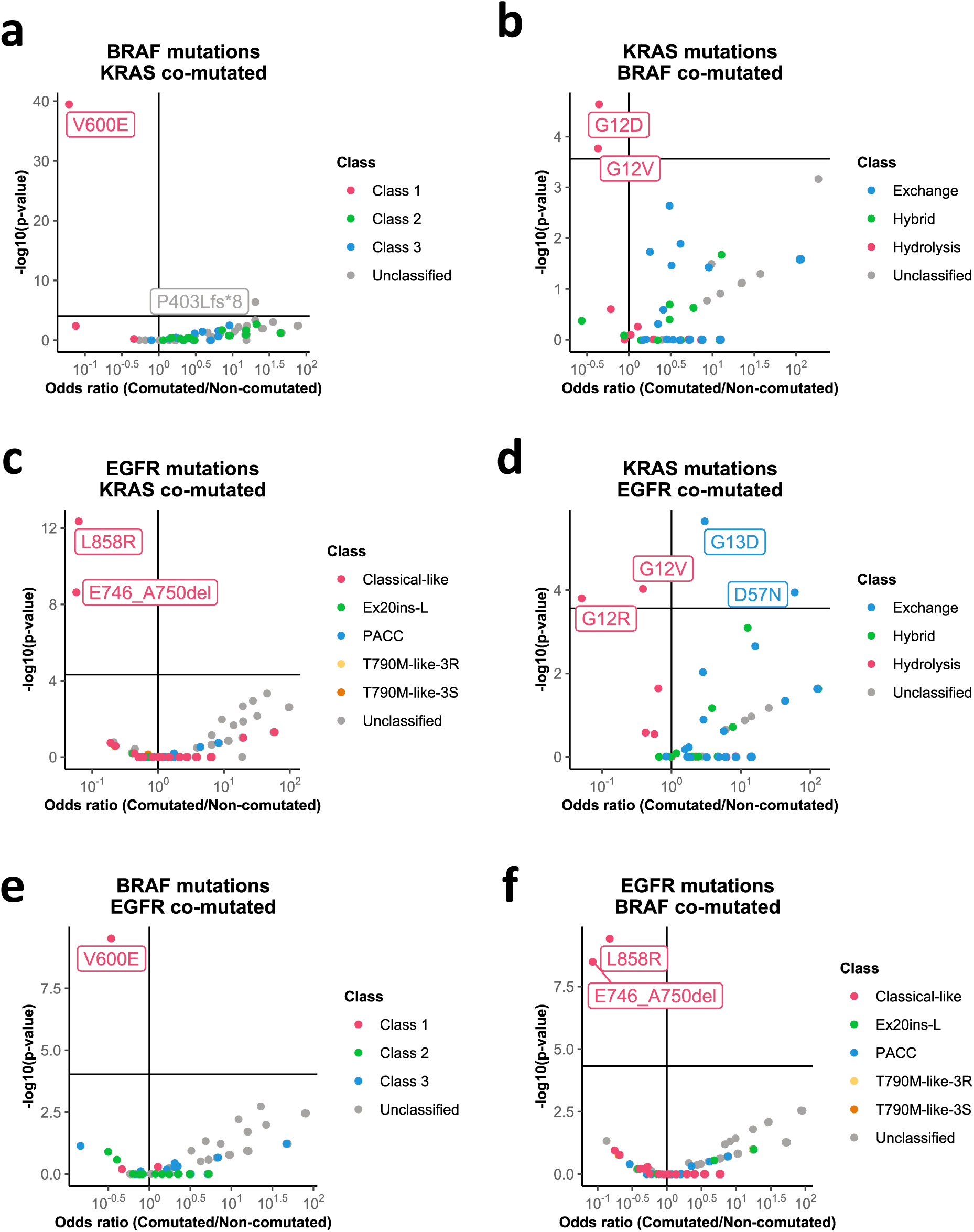
BRAF, KRAS, and EGFR mutations with Mutually Exclusive or Co-occurring tendencies. Using 2x2 contingency tables (as detailed in the methods section), we computed odds ratios (ORs) and two-sided Fisher’s Exact test p-values. The scatter plots display the ORs of the respective unilaterally class-assigned variants corresponding to the CO group divided by the ME group against the negative decadic logarithm of the p-values. The horizontal line borders the statistical significance based on the Bonferroni-transformed p-value threshold, and the vertical line signifies the scenarios with equal frequencies in both CO and ME groups. As such, the up-left quarter of each plot represents the mutations that are significantly ME with the comparing genes, and the up-right quarter displays variants that are significantly co-mutated with the comparing genes. Note that the comparing genes are not class-assigned, e.g., BRAF^V600E^ with KRAS mutations reported in cancer, irrespective of KRAS class. (**a**) shows the class-assigned BRAF variants occurring alone or in BRAF∩KRAS. (**b**) class-assigned KRAS variants alone or in BRAF∩KRAS. (**c**) class-assigned EGFR variants alone or in EGFR∩KRAS. (**d**) class-assigned KRAS variants alone or in EGFR∩KRAS. (**e**) class-assigned BRAF variants alone or in BRAF∩EGFR. (**f**) class-assigned EGFR variants alone or in BRAF∩EGFR. Class assignment of the specific variants is indicated by the color code.

**Table. 2a:** Co-occurrence or mutual exclusivity among unilaterally class-assigned BRAF, KRAS, and EGFR gene variants found in human cancer samples. Two-sided Fisher’s Exact Test and the 2 × 2 contingency tables, as described in the methods, were used to address the co-occurrence or mutual exclusivity of class-assigned gene variants versus the comparing gene mutations compiled as one entity. The analyses were performed on publicly available data extracted via cBioportal, as in Figure 1. **Co Group** represents the co-occurring gene pairs, e.g., BRAF class I, co-occurring with KRAS mutations reported in human cancer, irrespective of classification for KRAS mutations. **Class** refers to classifications described in Table 1. Each Co Group is broken down into two class sections, each corresponding to one of the gene components in the specified gene pair. **(N) Class in co** communicates the number of samples that belong to the specified class in the specified co-occurring gene pair group. **(N) non-Class in co** represents the number of samples excluding the ones belonging to the specified class in the specified co-occurring gene pair group. **(N) class in non-co** displays the number of samples belonging to the specified class when mutually exclusive with the specified component in the gene pair. **(N) non-class in non-co** communicates the number of samples that do not belong to the specified class and are found in the mutually exclusive subset concerning the specified gene pair. Based on 2x2 contingency tables described in the methods, **Two-sided Fisher’s** *p-value*s were calculated and displayed. After the Bonferroni adjustment of the significance threshold, whether the **Two-Sided** test result is **Significant** was determined. The **Odds Ratio** and the standard error (**SE**) are also displayed as inferred.

| Co Group | Class | (N) Class in<br>co | (N) non-Class<br>in co | (N) class in<br>non-co | (N) non-class in<br>non-co | Two-sided Fisher's<br>*p-value* | significant<br>Two-Sided | Odds Ratio | SE |
| --- | --- | --- | --- | --- | --- | --- | --- | --- | --- |
| **BRAF∩KRAS** | **BRAF class** |  |  |  |  |  |  |  |  |
|  | I | 12 | 156 | 1558 | 983 | 5.89E-47 | TRUE | 0.05 | 0.30 |
|  | II | 28 | 140 | 191 | 2350 | 1.80E-04 | TRUE | 2.49 | 0.22 |
|  | III | 28 | 140 | 223 | 2318 | 1.45E-03 | TRUE | 2.10 | 0.22 |
|  | **KRAS class** |  |  |  |  |  |  |  |  |
|  | Hydrolysis | 82 | 86 | 4811 | 1463 | 1.35E-14 | TRUE | 0.29 | 0.16 |
|  | Exchange | 54 | 114 | 946 | 5328 | 5.46E-08 | TRUE | 2.68 | 0.17 |
|  | Hybrid | 13 | 155 | 422 | 5852 | 5.36E-01 | FALSE | 1.20 | 0.29 |
| **EGFR∩KRAS** | **EGFR class** |  |  |  |  |  |  |  |  |
|  | Classical_like | 8 | 140 | 1363 | 1486 | 5.81E-29 | TRUE | 0.07 | 0.36 |
|  | Ex20ins_L | 0 | 148 | 50 | 2799 | 1.76E-01 | FALSE | 0.19 | 1.42 |
|  | PACC | 3 | 145 | 164 | 2685 | 6.30E-02 | FALSE | 0.39 | 0.55 |
|  | T790M_like_3R | 0 | 148 | 3 | 2846 | 1.00E+00 | FALSE | 2.74 | 1.51 |
|  | T790M_like_3S | 1 | 147 | 111 | 2738 | 4.24E-02 | FALSE | 0.25 | 0.83 |
|  | **KRAS class** |  |  |  |  |  |  |  |  |
|  | Hydrolysis | 66 | 82 | 4824 | 1466 | 1.79E-16 | TRUE | 0.25 | 0.17 |
|  | Exchange | 54 | 94 | 946 | 5344 | 2.36E-10 | TRUE | 3.26 | 0.17 |
|  | Hybrid | 15 | 133 | 420 | 5870 | 9.81E-02 | FALSE | 1.62 | 0.27 |
| **BRAF∩EGFR** | **BRAF class** |  |  |  |  |  |  |  |  |
|  | I | 55 | 106 | 1514 | 1033 | 5.53E-10 | TRUE | 0.36 | 0.17 |
|  | II | 3 | 158 | 215 | 2332 | 1.39E-03 | TRUE | 0.24 | 0.55 |
|  | III | 11 | 150 | 240 | 2307 | 3.27E-01 | FALSE | 0.73 | 0.31 |
|  | **EGFR class** |  |  |  |  |  |  |  |  |
|  | Classical_like | 9 | 152 | 1362 | 1474 | 3.92E-31 | TRUE | 0.07 | 0.34 |
|  | Ex20ins_L | 2 | 159 | 48 | 2788 | 1.00E+00 | FALSE | 0.90 | 0.65 |
|  | PACC | 5 | 156 | 162 | 2674 | 2.14E-01 | FALSE | 0.58 | 0.44 |
|  | T790M_like_3R | 1 | 160 | 2 | 2834 | 1.53E-01 | FALSE | 10.60 | 1.04 |
|  | T790M_like_3S | 0 | 161 | 112 | 2724 | 4.09E-03 | FALSE | 0.07 | 1.42 |

*BRAF∩KRAS -* class I BRAF is significantly less frequent in the CO group, and class I variant V600E is ME with KRAS mutations (Table 2a and Fig. 2a). In line with a recent report by Zhao et.al^7^, we found a higher proportion of atypical BRAF mutations, both class II and III BRAF, in the KRAS∩BRAF group.

Unlike Zhao et.al^7^, besides BRAF, we also acknowledge KRAS classes in our analysis. Interestingly, we discovered that the Hydrolysis KRAS class is ME with BRAF mutations (Table 2a). In particular, KRAS^G12D/V^ are significantly ME with BRAF mutations (Fig. 2b). On the contrary, the Exchange KRAS class is enriched in the CO group. Interestingly, minimally characterized BRAF^P403Lfs*8^ was the only BRAF variant that showed statistical significance in its co-occurrence with KRAS mutations. This variant is suggested to be a passenger mutation known to arise due to high tumor mutational burden in mismatch repair deficient tumors^25^. The variant has also shown a growth inhibitory effect when overexpressed in Ba/F3 and MCF10A cells^26^.

*EGFR∩KRAS -* among EGFR classes, the Classical-like class, which includes the predominant EX19Del- and L858R mutations, is ME with KRAS mutations (Table 2a). EGFR^L858R^ and EGFR^E746_A750Del^ are the only significant ME variants with KRAS mutations (Fig. 2c). The Hydrolysis KRAS class is ME with EGFR mutations, while the Exchange KRAS class is enriched in the CO group (Table 2a), with KRAS^G12R/V^ (Hydrolysis) being significant in their mutual exclusivity (Fig. 2d). KRAS^G13D^ and KRAS^D57N^ (both Exchange) are significantly enriched in EGFR∩KRAS (Fig. 2d).

*BRAF∩EGFR -* we found both class I and class II BRAF, but not class III are ME with EGFR mutations (Table 2a). The Classical-like EGFR class is ME with BRAF mutations. T790M-like-3S showed only a statistically insignificant trend of mutual exclusivity.

BRAF^V600E^, EGFR^L858R,^ and EGFR^E746_A750Del^ are significantly ME with EGFR and BRAF mutations, respectively (Fig. 2e&f).

Overall, excluding the type I BRAF and in particular BRAF^V600E^, different gene classes showed heterogenous trends of ME and CO with other gene variants.

### Analyses of Cancer cell line database affirms Mutually Exclusive scenarios concluded in patients’ database

Next, we sought to determine whether similar conclusions could be drawn in cancer cell lines, as in patients’ tumor samples. We queried 1570 mutant cell line data, available in Cancer Cell Line Encyclopedia (Broad, 2019), and considered the ones with BRAF, KRAS, or EGFR mutations. We carried out an analysis identical to the one performed on patient data to address the distinct proportions of mutational classes within cancer cell lines (Table 2b). Compared to patients’ tumor samples, we did not find an increased frequency of KRAS Exchange or BRAF class III variants in any CO scenarios. Due to the relatively limited number of available cell lines with CO scenarios, robust statistical conclusions could not be drawn. The only ME scenario that was statistically significant was between class I BRAF and KRAS mutations. Still, when comparing the ORs of patients and cell line CO scenarios, 17/22 showed a similar trend (Table 2b).

**Table. 2b:**
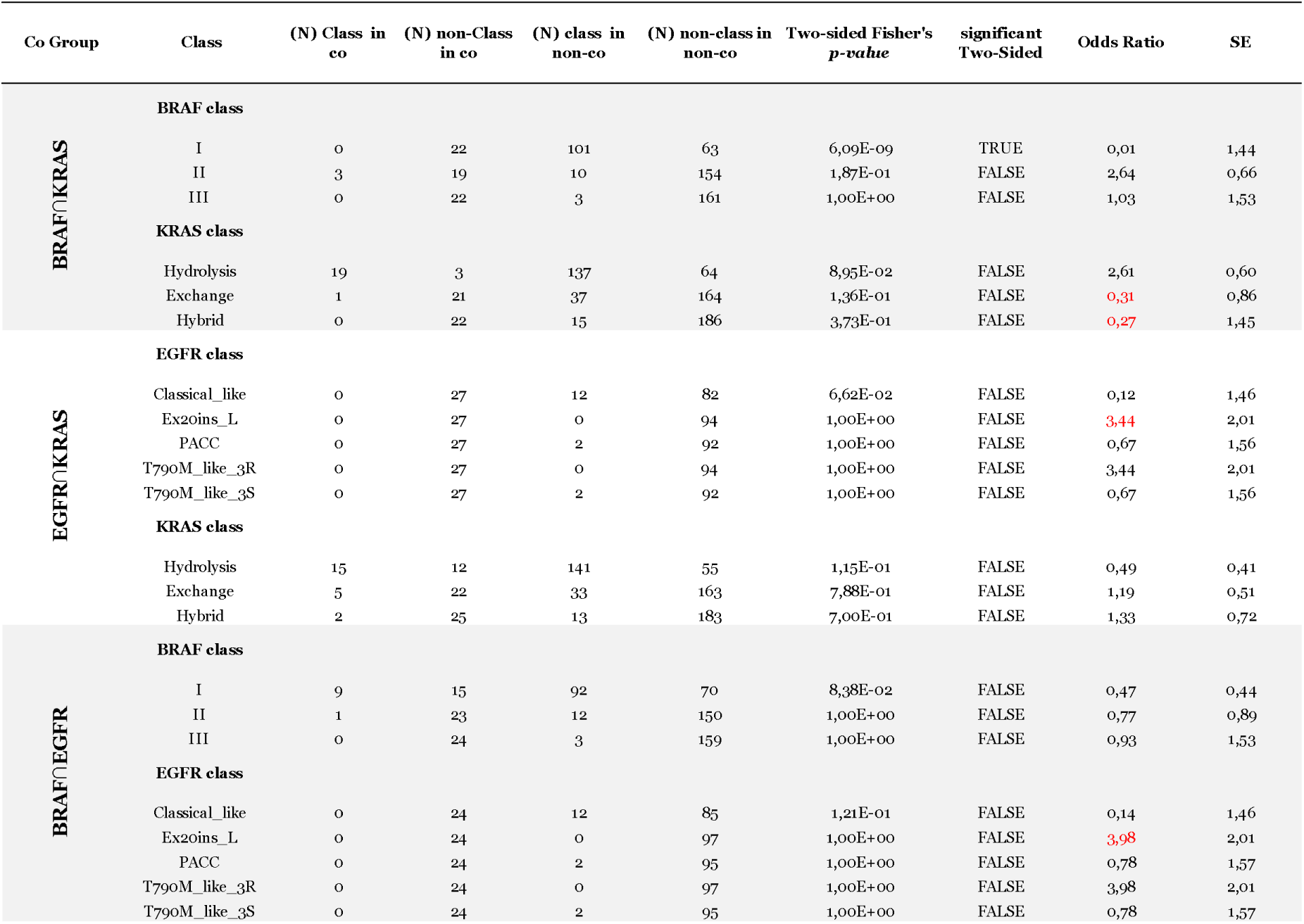
Co-occurrence or mutual exclusivity among unilaterally class-assigned BRAF, KRAS, and EGFR gene variants found in human cancer cell lines. The same analyses as in Table 2a but on 1570 mutant cell line data, available in Cancer Cell Line Encyclopedia (Broad, 2019), and considering the ones with BRAF, KRAS, or EGFR mutations (see Figure 1a). When ORs do not follow the same trend as in patients’ samples, they are mentioned in red. Class I and III BRAFs were absent in BRAF∩KRAS cell lines.

So far, among the CO cell lines, we did not find any cell line with concomitant mutations belonging to BRAF class I, KRAS Hydrolysis class, or EGFR Classical-like class. As such, our analysis of the cell line database is affirmatory concerning the significant ME trends among the patients.

### Unraveling the co-occurrence landscape of BRAF, KRAS, and EGFR mutations in human cancer

Thus far, we have examined the relative representation of BRAF, EGFR, and KRAS mutational classes and types in CO scenarios as compared to non-CO situations. This approach may not definitively determine whether co-occurring scenarios involve driver mutations of established significance or if the pairs might include passenger variants or those with unknown functional impact. Yet, we were interested in addressing the inter-class and inter-type co-occurrence and mutual exclusivity patterns. Hence, we generated co-occurrence heatmaps for each of the three CO scenarios, considering the absolute frequency of each variant. For this analysis, we included only the variants that were observed at least five times in human cancer (Fig. 3).

**Fig. 3:**
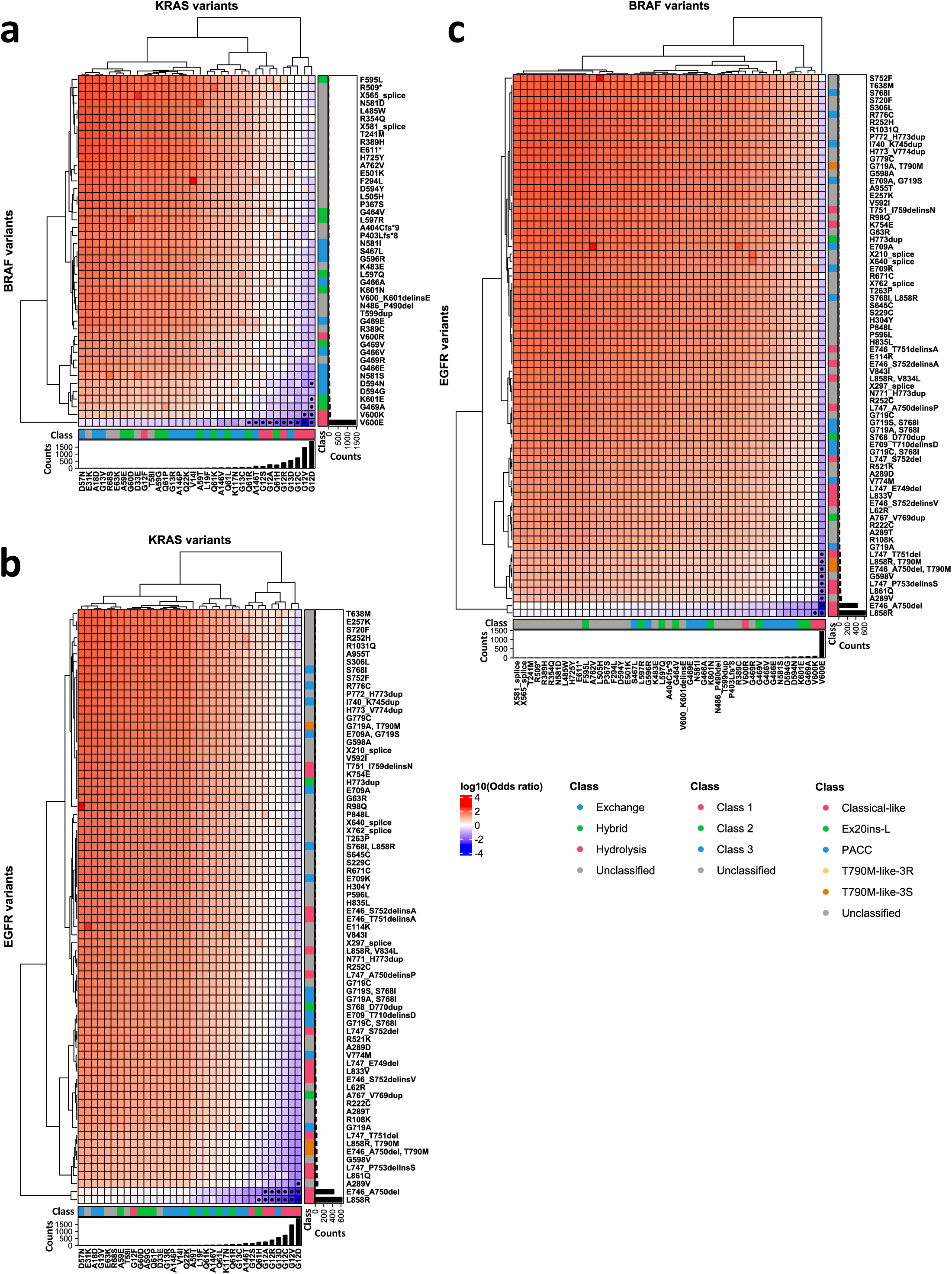
The co-occurrence landscape of BRAF, KRAS, and EGFR mutations in human cancer. Heatmaps representing co-occurrence of reported variants of (**a**) KRAS and BRAF, (**b**) KRAS and EGFR or (**c**) BRAF and EGFR in human cancer observed in at least five patient samples. Log10 odds ratios range from -4 (deep blue, most mutually exclusive) to 4 (deep red, highest co-occurrence), with white squares indicating equal frequencies in ME and CO. Colored bands on axes indicate gene classes, and black bars represent absolute variant frequencies. Dots denote statistical significance. The related tables can be found in Supplementary Table 1-3.

In *BRAF∩KRAS* (Fig. 3a), as for the other two CO groups (Fig. 3b&c), mutual exclusivity was not an exclusive feature of any class (Fig. 3a). Interestingly, the most frequent mutations, such as BRAF^V600E^ and KRAS^G12D,^ were the most ME ones, as indicated by the lower values of the Odds ratio (OR). These statistically significant ME scenarios encompassed certain members of all three BRAF and KRAS classes (Fig. 3a). Conversely, variant rarity was linked to co-occurrence, as evidenced by a relatively higher OR among the less frequent mutations. Yet, the highly CO scenarios were not statistically significant.

In *EGFR∩KRAS,* as expected, EGFR^EX19Del^ and EGFR^L858R^ mutations were among the most mutually exclusive variants with predominant KRAS variants (Fig. 3b). Like BRAF∩KRAS, all three KRAS classes could be represented by their certain members among the variants with the highest ME score. Among the classified EGFR variants, only Classical-like was represented in highly ME scenarios. Interestingly, EGFR^A289V^, an atypical variant (when occurring alone but not with L858R), was among the EGFRs with the highest and statistically significant ME score.

In *BRAF∩EGFR*, we identified specific instances of Classical-like, T790M-like-3S, or unclassified EGFR mutations (EGFR^G598V^ and EGFR^A289V^) to be significantly ME with BRAF^V600E^ (Fig. 3c).

Overall, our co-occurrence matrices revealed a few novel ME scenarios. From a broader perspective, we unraveled the pan-cancer mutual exclusivity landscape among BRAF, KRAS, and EGFR mutations. The link between mutational frequency and mutual exclusivity is a hallmark of this landscape.

Next, we sought to pinpoint the samples and the corresponding tissue types where these extreme ME scenarios still co-occurred. Curiously, only nine samples harbored such exceptional pairs. Interestingly, among six samples with existing information on the allele frequency (AF) for each oncogene in a tumor, four samples had an AF for one of the co-occurring components (KRAS) smaller than 10 percent, suggesting a possibility that these mutations might not be co-occurring in the identical tumor compartments (see Table 3) or might be the result of contamination. Moreover, in line with a previously reported phenomenon of secondary resistance to EGFR-targeted therapies^12,13^, the only BRAF∩EGFR sample (Table 3) was derived from a patient under anti-EGFR therapy^27^.

**Table. 3:**
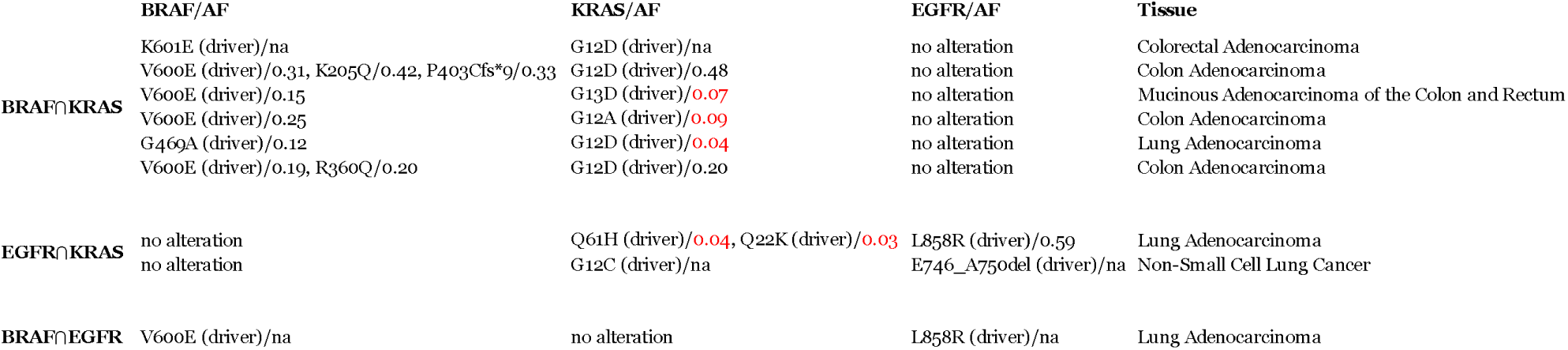
Samples with yet co-occurrence of ME scenarios with statistical significance and their related tissue types. **AF**: denotes Allele Frequency in the tumor, **na**: not available. AFs <0.1 are mentioned in red. Note that non-small lung cancer and colorectal cancer account for the 2^nd^ and 3^rd^ most frequent samples in our queried dataset after breast cancer (see supplementary figure 1). These two cancer types are known to harbor KRAS, BRAF, or EGFR mutations recurrently. We calculated binomial probabilities encountering exactly these cancer types upon random sampling. The likelihood of BRAF∩EGFR occurring exactly 1 out of 1 time in Non-Small Cell Lung Cancer was calculated as p = 0.1010, CI = (0.03, 1.00), the same for the following was inferred: KRAS∩BRAF 5 of 6 of Colorectal Cancer p = 0.0000, CI = (0.36, 1.00), KRAS∩BRAF 1 of 6 of Non-Small Cell Lung Cancer p = 0.4721, CI = (0.00, 0.64) and the KRAS∩EGFR 2 of 2 of Non-Small Cell Lung Cancer p = 0.0102, CI = (0.16, 1.00). Note that confidence intervals for these p-values are so large, hindering any robust statistical conclusions.

### Mutual Exclusivity and Co-occurrence in a Belgian cancer patients’ dataset

As observed, mutational classifications exhibit high variability in terms of CO trends among different variants. Thus far, our findings underscore the significance of the variant-specific landscape of ME. In the cBioPortal database, variant allele frequencies in the tumor are not available for all samples, and we did not acknowledge the corresponding data in our analysis unless in exceptional ME scenarios. Additionally, we included all the reported mutations in our previous analysis regardless of existing estimations of their impact.

As such, we aimed to validate our findings in a manually curated dataset of 2714 samples from Belgian cancer patients who underwent targeted NGS analysis at UZ-Brussel Hospital.

An Upset plot (Fig. 4a) illustrates the distribution of BRAF, KRAS, and EGFR variants among six tumor types (1711 samples), with frequencies consistent to prior findings in the literature. Within this constrained dataset, the occurrence of variants in gene pairs is concentrated within specific tumor types as further detailed in the figure’s legend.

**Fig. 4:**
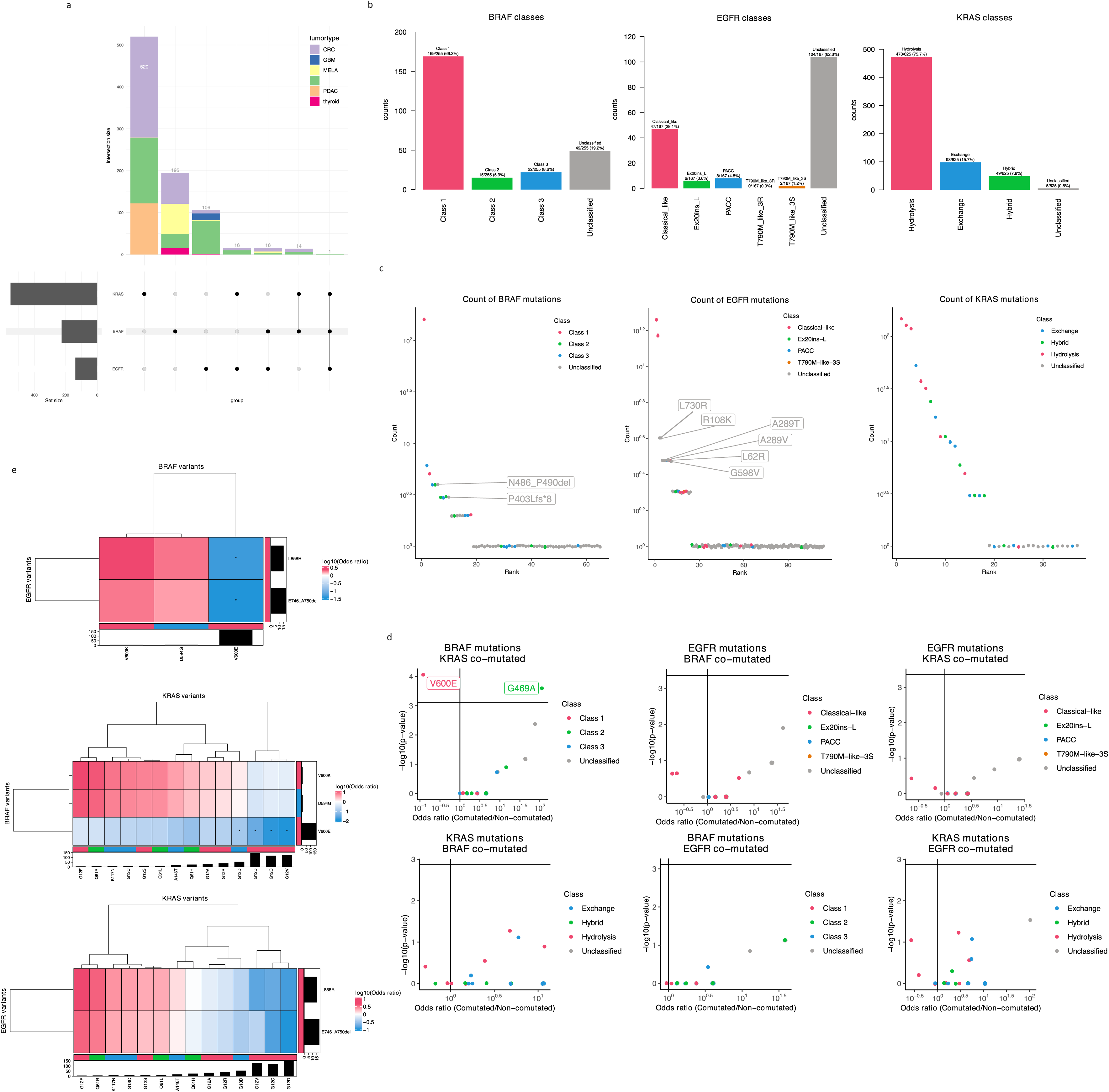
Mutual Exclusivity and Co-occurrence in a Belgian cancer patients’ dataset. The Belgian cancer NGS dataset encompasses curated variants of 2714 unique samples from patients with solid tumor types across several tissue types, collected between July 01, 2019, and April 24, 2023. The dataset includes 260 BRAF, 636 KRAS, and 184 EGFR variants (SNV’s, splice site variants, and short indels), of which, respectively, 63, 37, and 115 are unique gene variants. In (**a**), an Upset plot displays the distribution and overlap of 231 BRAF, 568 KRAS and 154 EGFR-variants in 1711 patient samples among six tumor types, each harbouring at least 5% of all BRAF, KRAS or EGFR variants identified in the complete Belgian dataset (NSCLC; Non-Small Cell Lung Cancer, CRC; Colorectal cancer; PDAC; Pancreatic Ductal Adenocarcinoma; MELA; Melanoma, GBM; Glioblastoma and Thyroid cancer). Variants are listed in supplementary Table 5. The frequencies of gene-specific variants in the six tumor types align with previously reported evidence in the literature. The CRCs, NSCLCs, and PDACs are recurrently found to harbor KRAS variants^38^. In NSCLC, this is followed by EGFR variants and subsequently BRAF variants. In CRC, however, this order is reversed^3,4^. BRAF variants are typically associated with thyroid cancer and melanoma^3^. In glioblastomas, EGFR is often affected by variants in the extracellular region of the protein^39^. The occurrence of variants in specific gene pairs is mainly restricted to NSCLC and CRC; BRAF∩KRAS in six NSCLCs and eight CRCs, BRAF∩EGFR in four NSCLCs, nine CRCs, and three melanomas, KRAS∩EGFR in nine NSCLCs, six CRCs, and one endometrial carcinoma (not shown in Fig. 4a). One NSCLC tumor was found with coexistence of mutations in all three mentioned genes. In (**b**), bar charts display the relative frequency of BRAF, KRAS, and EGFR mutational classes as percent in all 2714 tested Belgian cancer samples. These frequencies closely align with those identified in the cBioPortal patient dataset, apart from the EGFR class distribution. For EGFR, we found fewer Classical-like variants (28.1% vs. 45.4%) but a higher frequency of unclassified variants (62.3% vs. 43.6%). One possible explanation for this variation could be attributed to the assumed composition of the Belgian test population, which is expected to include a smaller proportion of patients with Asian ethnicity. However, it should be noted that no demographic data were included in the analysis. The Belgian dataset also encompassed glioblastoma cases, which often feature EGFR mutations within the extracellular domain, probably contributing to the slightly increased occurrence of unclassified EGFR variants (3% Belgian GBM vs. 1% cBioPortal GBM)^39^. In sections (**c**), (**d**) and (**e**), we provide graphs, scatter plots and heatmaps showcasing the entire Belgian dataset and generated analogously as for Figure 1b, Figure 2 and Figure 3, respectively. In (**c**), several unclassified variants are among the top 10 variants. For BRAF these include BRAF^N486_P490del^ and BRAF^P403Lfs*8^, and for EGFR they include EGFR^A289V^, EGFR^A289T^, EGFR^G598V^, EGFR^R108K^, and EGF^RL62R^ (all recurrent in GBM) and also EGFR^L730R^ (primarily detected in NSCLC). In (**d**), we exclusively observe significant senarios involving KRAS mutations, such as with Class I BRAF^V600E^ in ME pattern and with BRAF^G469A^ in CO pattern. In (**e**), the co-occurrence heatmaps indicate CO scenarios involving less frequent mutations (without statistical significance) while ME senarios do not correspond to any specific class but exhibit significance for specific mutation types.

Bar charts (Fig. 4b) illustrate the class frequencies of BRAF, KRAS, and EGFR genes within the complete Belgian dataset. The figure’s legend highlights the distinctive frequencies of each class compared to the cBioPortal samples. Few unclassified BRAF variants are within the top ten variants, including BRAF^N486_P490del^ followed by BRAF^P403Lfs*8^ (Fig. 4c). The specific BRAF^P403Lfs*8^ variant, statistically significant in CO with KRAS mutations in the cBioPortal dataset, is found in three Belgian cases, once within an adrenocortical carcinoma harbouring KRAS^A146T^ and twice within the context of mismatch repair deficiency (CRC and endometrial carcinoma). Several other atypical variants are also in the top ten frequent EGFR variants list. These include EGFR^A289V^, EGFR^A289T^, EGFR^G598V^, EGFR^R108K^, and EGFR^L62R^, which exhibit a recurring pattern in glioblastoma cases, whereas EGFR^L730R^ is predominantly found in NSCLC.

For the Belgian dataset, the relation of class-assigned gene variants to not-class-assigned variants of the compared genes, is presented in Figure 4d. Among the EGFR classes, no significant patterns in CO or ME with KRAS or BRAF variants were identified. The same applied to KRAS classes and BRAF classes, each in relation to EGFR variants. Class I BRAF^V600E^ follows the significant trend in ME with KRAS mutations, while class II BRAF^G469A^ is significant in CO with KRAS mutations. However, we should consider that we found only three samples with BRAF^G469A^ in our dataset. Therefore, no solid conclusion can be drawn in this case. The BRAF^G469A^ co-occurred with KRAS^G12V^, KRAS^G12C^ (both CRC) and KRAS^G12A^ (NSCLC). Interestingly, as we observed in statistically significant ME cases in Figure 3a, none of these co-occurring variants involved the KRAS^G12D^.

Finally, co-occurrence heatmaps (Fig. 4e) were generated for each of the three CO scenarios in the Belgian dataset. Similar to the cBioPortal data, CO scenarios occur only with less frequent mutations being not statistically significant, and ME does not align with any specific class. For EGFR∩KRAS and EGFR∩BRAF, we confirm that Classical-like variants EGFR^EX19Del^ and EGFR^L858R^ are both ME with respectively Hydrolysis KRAS^G12V/D/C^ variants (not statistically significant) and class I BRAF^V600E^ (statistically significant). In KRAS∩BRAF, we identified class I BRAF^V600E^ to be significantly ME with Hydrolysis KRAS^G12V/D/C^ variants as well as with the Exchange KRAS^G13D^ variant.

Overall, in line with our prior analysis, gene classifications exhibited significant variability in terms of CO patterns with other gene variants. In this regard, disparities between our findings in the cBioPortal analysis and the Belgian cancer dataset were observed. Once again, we observed that a variant-specific landscape may offer more informative insights into co-occurrence and mutual exclusivity patterns than the gene classifications.

## Discussion

Overall, Type I BRAF, Hydrolysis KRAS, and Classical-like EGFR class mutations, all known as strong activators of the ERK pathway, are less likely to co-occur with each other. ^3–5^. Of note, our co-occurrence heatmaps with bilateral class/variant assignments revealed several novel ME scenarios. For instance, the first part of our analyses concluded that BRAF class II is enriched in the BRAF∩KRAS group, in line with a recent report by Zhao et al.^7^. However, heatmaps enabled the discovery of mutual exclusivity of the well-studied class II BRAF^G469A^ and KRAS^G12D^. Additionally, we discovered some previously unknown ME pairs encompassing atypical EGFR mutations with BRAF and KRAS mutations that warrant further preclinical investigations.The ME scenarios showed statistical significance as opposed to co-occurring pairs. In a pan-cancer analysis of mutual exclusivity among somatic mutations, Canisius et al. propose that mutual exclusivity in cancer is a product of active biological phenomena, while co-occurrences are somewhat random^28^. As such, our findings could be in line with Canisius et al. conclusions. Conversely, the absence of statistical significance could be attributed to such scenarios’ limited frequency. Furthermore, it is crucial to consider an inherent limitation of pan-cancer analyses, including our study and the work by Canisius et al., where the significance of context-dependent associations might be overlooked as all tissue types are conflated within a singular analytical framework.

One could not determine that simply the redundancy of function lies behind the mutual exclusivity of some mutations, or rather, an active biological phenomenon could limit or even exclude such co-occurring events. To tailor our revisit of the precedent literature to the ME scenarios in our current findings, we systematically reviewed the literature for studies that have modeled concomitant induction of the very oncogenic variants in the same cell, as described in this study (supplementary table 6). Briefly, in different preclinical lung cancer models, co-expression of BRAF^V600E^∩KRAS^G12D^ ^8^, KRAS^G12V^∩EGFR^L858R^ ^29^, EGFR^EX19Del^∩KRAS^G12C^ ^29^, and EGFR^L858R^∩KRAS^G12V^ ^9^ has been investigated. Across all these studies, the co-induction of two oncogenes could not be tolerated in the same cell. The coexistence of both oncogenes was synthetically lethal or led to senescence^8,9,29^. In these cases, the observed phenomenon was at least partly attributed to supra-physiological levels of MAPK signaling^8,9,29^. On the other hand, we did not find direct biological evidence relating the mutual exclusivity of oncogenic BRAF and EGFR mutations to senescence or synthetic lethality. So far, two studies have been inspired by the emergence of BRAF^V600E^ in EGFR^L858R^ relapsed lung cancers as they become resistant to EGFR-targeted therapies^12,13^. Therefore, as the precedent literature suggests, active biological phenomena such as senescence and synthetic lethality can limit the possibility of few of the CO events described in this study, at least in BRAF∩KRAS and EGFR∩KRAS pairs. One should consider that redundancy of functions and synthetic lethality are not necessarily conflicting scenarios, as the latter can involve excessive activity of two redundant functions. As such, they might both contribute to the rise of mutual exclusivity. Yet, further benchwork and collective effort are essential to unveil the mechanisms behind the novel ME pairs reported in this study.

Based on the available patient dataset, we could not determine whether BRAF, KRAS, and EGFR mutations had occurred in the same cells in CO groups. Yet, our analysis of cell lines could address this question. Interestingly, in a recent study^30^ benefiting from single-cell RNA sequencing of tumor samples, the authors report that in a cohort of treatment-naive Colorectal Cancer patients, albeit at a very low frequency, BRAF^V600E^ and KRAS^G12D^ events could exist in the same cell. Markedly, this co-occurrence was accompanied by the compromise of the allelic imbalance, a phenomenon that favors the oncogenes’ dose increase by different mechanisms in cells with a single oncogene (RAS or BRAF). Once again, these findings align with the concept of *oncogene overdose*^31^. In general, human cells, including cancer cells, cannot exceed a certain threshold of ERK pathway activity^32–34^.

Our CO matrices within the Belgian dataset confirmed the ME scenarios, provided that both variants had previously been included in the analysis of the sample data available in cBioPortal. As we further observed in the Belgian cancer patient dataset, all three instances of BRAF^G469A^ occurrence coincided with KRAS activating variants. This contradicts prior conclusions that has classified BRAF^G469A^ as RAS-independent^3^. It is worth mentioning that none of the co-occurring KRAS variants were KRAS^G12D^. This underscores the robustness of our approach in revealing variant-specific mutual exclusivity in human cancer. Yet, it is important to remember that these ME scenarios represent tendencies rather than absolute phenomena. Furthermore, we highlight several EGFR variants that had previously been overlooked in terms of their significance. These variants were found to be recurring in both publicly available and our clinical dataset.

Apart from the mutation types, we did not address the role of other players that may impact the mutual inclusivity or exclusivity among BRAF, EGFR, and KRAS mutations. Such players could include effectors and regulators of ERK, their status in cancer, and other potential confounders, such as tissue context, age, sex, treatment, and disease stage. Regarding some variants, such as EGFREx20ins-L and PACC class members, our consulting dataset was limited, hindering solid statistical conclusions. Moreover, we did not address the chronological order of gene mutations in patients with CO gene pairs.

Our findings on classes with diverse co-occurrence patterns emphasize the need to revise the mutational classifications for BRAF, KRAS, and EGFR. In CO mutation scenarios, context- and mutual dependencies should be investigated. The mechanisms elicited by the coincidence of ME events described in this study deserve to be investigated as they can guide us through unexplored vulnerabilities and novel therapeutic opportunities in ERK-related cancers.

## Methods

### Pilot analysis

Initially, TSV tables of the BRAF-, KRAS- and EGFR-mutant patient samples and cell lines were extracted from cBioportal. Class proportions were assessed in Microsoft Excel 2010 to determine the frequency of mutation types as well as mutation classes. Fisher’s exact test was performed in GraphPad prism to compare mutations class proportions between comparing groups.

### R Code

The R scripts employed for conducting analyses, generating figures, and creating tables in this study, along with detailed explanations, are readily accessible to the public (please consult the data availability section). The specific version utilized for this work is R version 4.2.3 (2023-03-15 ucrt). ComplexHeatmap package was utilized to visualize the heatmaps^35,36^.

### Source of Data

Patient data with information about the mutation status of KRAS, BRAF, and EGFR genes from 213 curated and non-redundant studies were obtained from cBioPortal. The dataset was up to date as of November 15, 2022, and encompassed details from a total of 68479 samples, representing 64911 patients. After removing duplicate patients determined by cross-referencing the patient I.D. and excluding multiple samples from the same patient while retaining the first sample collected, 64807 patient data remained for further analysis. For the analysis of the mutation status of different cell lines, the mutation data of 1570 different cell lines from the Cancer Cell Line Encyclopedia (Broad, 2019) study of cBioPortal was queried. The dataset was current as of December 01, 2022.

### Class assignment

Patients were categorized into mutation classes for KRAS, BRAF, and EGFR genes using established tables derived from the most recent mutational classifications^3–5^. Single mutations were assigned to their respective mutation classes based on patient data analysis. Patients with multiple class-defining mutations were assigned to all corresponding classes, but each class was assigned only once per patient. Two special considerations applied to the classification of EGFR mutations. First, all deletions spanning amino acid 729 to 761 were considered as exon 19 deletions. Second, some class assignments were based on tuples of mutations rather than single mutations. Therefore, a specific rule was applied: if a patient had a mutation/tuple corresponding to a class assignment and also possessed all mutations of another tuple, the class assignment was based on the tuple with the larger number of mutations (e.g., if a patient had mutations L858R and T790M, they were assigned to the tuple “L858R, T790M” (T790M_like_3S) instead of L858R (Classical_like) and T790M (T790M_like_3S) alone. Note that this rule was introduced to assign the sample to the correct class as described in the respected reference.

### Statistical analysis for co-occurrence between mutation classes and mutated genes

For the analysis of the co-occurrence of mutation classes of gene G0 with the alteration status of gene G1, for each mutation class, a 2x2 contingency table for the presence of the corresponding mutation class of G0 and the alteration status of G1 was created from a subset encompassing all patients with mutated gene G0. Based on this 2x2 contingency table with elements A, B, C, and D, the Odds ratio (OR) was calculated according to the formula (A*D)/(B*C), the standard error (SE) according to the formula (1/A+1/B+1/C+1/D)^0.5^, and a p-value using two-sided Fisher’s Exact test. For contingency tables to address values of 0 for further calculations, we applied Haldane correction by adding 0.5 to each cell and then calculated OR and its SE. A p-value < 0.05 was considered statistically significant, but the p-values were adjusted for multiple testing according to the Bonferroni procedure. Concerning the subset consisting of all patients with mutated gene G0, the analysis was repeated for a subset composed of all patients with mutated gene G0 or with mutated gene G1. Statistical analyses were performed as described above.

### Statistical analysis for the co-occurrence of single mutations and altered genes

For the analysis of the co-occurrence of single mutations of gene G0 with the alteration status of gene G1, for each mutation, a 2x2 contingency table was created for the presence of the mutation of G0 and alteration status of G1 from a subset consisting of all patients with mutated gene G0. Utilizing this contingency table, which includes elements A, B, C, and D, we computed the OR, SE, and p-value following the procedures outlined earlier.

### Statistical analysis for co-occurrence between single mutations of two genes

To examine the co-occurrence of specific mutations within gene G0 and gene G1, we utilized a 2x2 contingency table approach. For each specific pair of mutations, we constructed a 2x2 contingency table within the defined subset that encompassed patients with either the mutated gene G0 or the mutated gene G1. This table assessed the presence of the corresponding mutation of G0 and the corresponding mutation of G1. Within this contingency table, featuring elements A, B, C, and D, we inferred the OR, SE, and p-value as outlined earlier. The statistical computations followed the methodologies detailed above.

### Procedure for constructing a curated Belgian cancer NGS dataset

In total, 2714 unique samples from patients with solid tumors across several tissue types were collected between July 01 2019, and April 24 2023. NGS was performed on FFPE-extracted DNA (Maxwell, Promega) using a BELAC accredited in-house capture-based comprehensive gene panel on Illumina NovaSeq6000 to reveal somatic variants. Subsequently an in-house developed script was run on all tested samples to examine for possible contaminations raised during the NGS wet lab procedure. Samples were flagged when more than 50% detected variants (<10% AF) were common with another sample (>30% AF) of the same NGS run. Corresponding variants of the flagged sample were only removed from the dataset after contamination confirmation in the lab. Finally, the dataset was limited to the genes BRAF, KRAS, and EGFR for which true variants were biologically and clinically classified according to the ComPerMed guidelines (https://www.compermed.be^37^). Variants only remained in the dataset if the biological impact was (likely) pathogenic or unknown. The dataset includes 260 BRAF, 636 KRAS, and 184 EGFR variants (SNVs, splice site variants, and short indels), of which, respectively, 63, 37, and 115 are unique gene variants.

## Data availability

The R script developed during this study can be found at https://github.com/HetzDra.

## Supplementary figures and table legends

**Supplementary figure.1:** Frequency of cancer types among the publicly available dataset queried for the current study.

**Supplementary Table. 1: Co-occurring Gene variants identified in the same cell line are listed here.** The only CO BRAF variants predicted to be drivers were BRAF^G464E/V^, belonging to class II. On the other hand, most of the KRAS mutations in this CO group belonged to the Hydrolysis class, and only one belonged to the Exchange class. Among EGFR∩KRAS cell lines, no classified EGFR mutation existed. EGFR^A864V^, EGFR^V292M^, and EGFR^A289V^ were the only variants predicted to be the driver. KRAS mutations in EGFR∩KRAS were more diverse than in BRAF∩KRAS, encompassing all three classes, but still dominated by Hydrolysis variants. Among BRAF∩EGFR cell lines, BRAF^V600E^ co-occurred only with non-driver/non-classified EGFR variants.

**Supplementary Table 2-4: Tables related to Figure 3**

**Supplementary Table. 5: Gene-specific variants identified in the Belgian dataset of six tumor types**

**Supplementary Table. 6: Precedent studies reveal that synthetic lethality and senescence lie behind mutual exclusivity among some oncogenic BRAF, KRAS, and EGFR events**

For each **study**, the investigated **Genes** variants and the study **Model** where the **Observed phenotype** was investigated are summarized. Unni et al., in a subsequent study, discovered DUSP6 as a signaling node that governs the observed phenotype^42^.

## Acknowledgments

Authors are grateful to all the institutions involved in this work.

## Funding

The work performed by the Centre for Human Genetics (Belgium) was co-funded by the Wetenschappelijk Fonds Willy Gepts of the UZ Brussel/VUB and the UZ Brussel Foundation.

Oleg Timofeev had received the following: The Federal Ministry of Education and Research (BMBF) grant 161L0279A and German Research Foundation (DFG) grant Ti1028/2-1.

Maxim Noeparast had received the following: Fonds Wetenschappelijk Onderzoek (FWO) -Vlaanderen, Belgium 12Y0120N.

## Author information

Authors and Affiliations

These first authors contributed equally: Freya Vaeyens and Jan-Patrick Hetzel

These corresponding authors contributed equally: Philippe Giron, Oleg Timofeev, and Maxim Noeparast

**Centre for Medical Genetics, Research Group Reproduction and Genetics, Clinical Sciences, UZ Brussel, Vrije Universiteit Brussel, Laarbeeklaan 101, 1090 Brussels, Belgium**

Philippe Giron, Ken Maes, Catharina Olsen, Freya Vaeyens, Jelle Vlaeminck and Frederik Hes

**Fonds Wetenschappelijk Onderzoek (FWO) -Vlaanderen, Belgium/ Laboratory for Medical & Molecular Oncology (LMMO)**

Maxim Noeparast and Carolien Eggermont

**Institute of Molecular Oncology, Member of the German Center for Lung Research (DZL), Philipps University, 35043 Marburg, Germany**

Jan-Patrick Hetzel, Marco Mernberger, Maxim Noeparast, Oleg Timofeev.

**Translational Oncology, University Medical Center Augsburg, Augsburg, Germany**

Maxim Noeparast and Martin Pichler

## Contributions

**Conceive**: MN **Conceptualization**: PG, OT, and M.N. **Pilot analysis**: MN, CE **Study and analyses design**: PG, OT, and MN with support from JPH **Supervision of the study**: PG, OT, and MN **Supervision on acquisition and curation of Belgian clinical data**: PG **Curation of Belgian cancer patients’ data**: FV as part of her Ph.D. work. **Collection and analysis of sequencing data**: CO, FV, JV, KM, PG **Statistical analyses** JPH and FV with support from CO and MM **Generation of figures and tables**: JPH, FV, and MN with support from MM, CO and CE **R codes**: JPH generated the codes, supplemented by CO and support from MM **Authoring the manuscript**: MN with shared contribution of FV and JPH **Critical revision of the text**: all co-authors.

## Ethics declarations

Ethical approval was obtained by the Medical Ethics Committee of UZ Brussel/VUB for studying NGS data from Belgian diagnostic cancer patients (EC-2023-040).

## Footnotes

♠ RTK/RAS/RAF (**R**eceptor **T**yrosine **K**inase/**Ra**t **S**arcoma/**R**apidly **A**ccelerated **F**ibrosarcoma), also known as MAPK or ERK pathway. In this writing, for simplicity, the pathway will be mentioned as ERK.

## Notes

### Competing Interest Statement

The authors have declared no competing interest.

