## Supplementary figures and images for "Variant-Specific Landscape of Mutual Exclusivity Among BRAF, EGFR, and KRAS Oncogenes in Human Cancer"

### Supplemental Figure 1

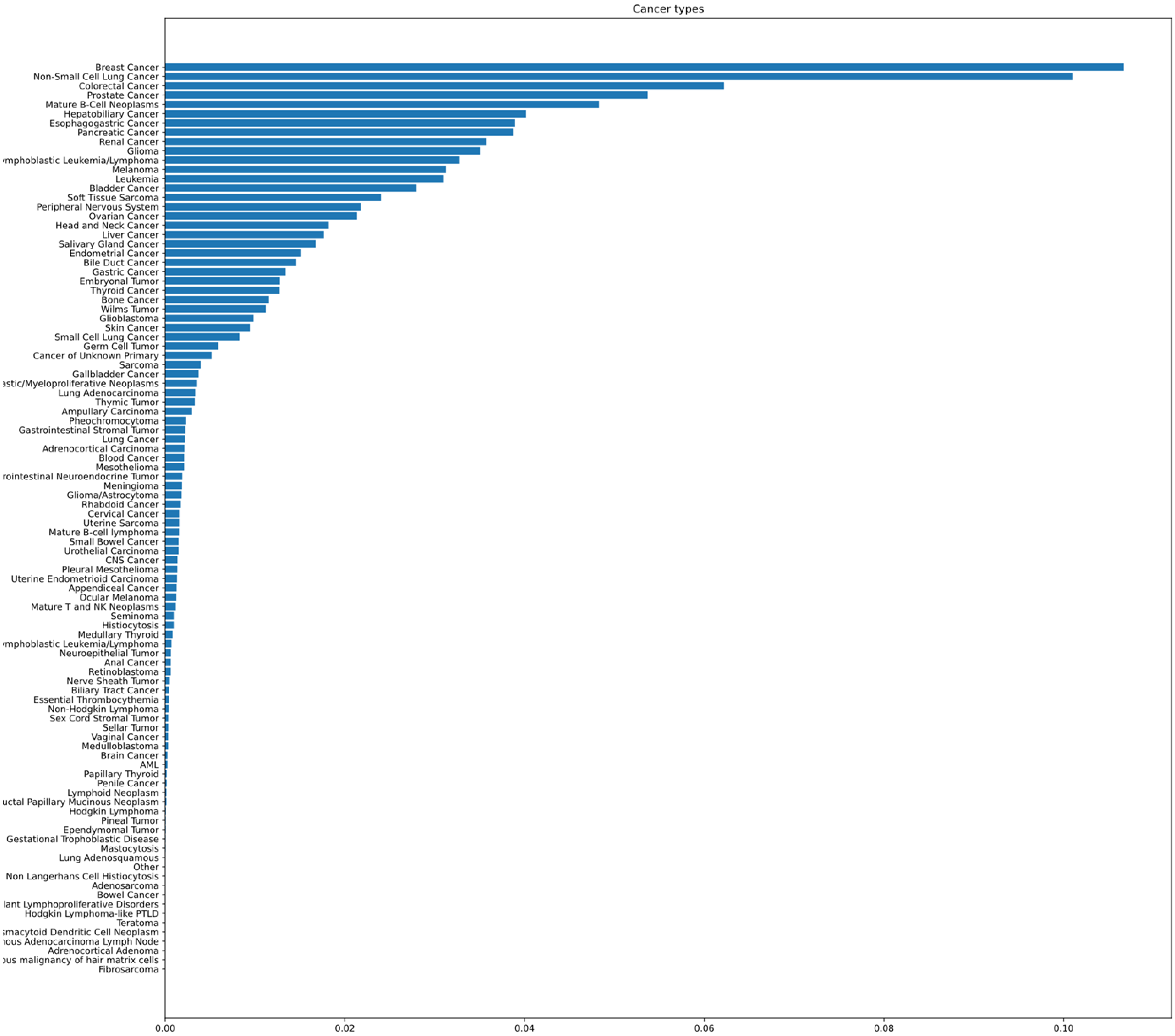
